# Searching for prognostic biomarkers of Parkinson’s Disease development in the Spanish EPIC cohort through a multiplatform metabolomics approach

**DOI:** 10.1101/2021.03.02.21252744

**Authors:** Carolina Gonzalez-Riano, Jorge Saiz, Coral Barbas, Alberto Bergareche, José Ma Huerta, Eva Ardanaz, Marcela Konjevod, Elisabet Mondragon, ME Erro, M. Dolores Chirlaque, Eunate Abilleira, Fernando Goñi-Irigoyen, Pilar Amiano

**Affiliations:** Centro de Metabolómica y Bioanálisis (CEMBIO), Facultad de Farmacia, Universidad San Pablo-CEU, CEU Universities, Urbanización Montepríncipe, Boadilla del Monte, 28660 Madrid, Spain; Neurodegenerative Disorders Area, Biodonostia Health Research Institute, San Sebastián, Spain; Disorders Unit, Department of Neurology, University Hospital Donostia, San Sebastián, Spain; Biomedical Research Networking Centre Consortium for the area of Neurodegenerative Diseases (CIBERNED), Madrid, Spain; Instituto Murciano de Investigación Biosanitaria (IMIB); CIBER de Epidemiología y Salud Pública (CIBERESP), Madrid, Spain; Instituto de Salud Pública de Navarra, Pamplona, Spain; Rudjer Boskovic Institute, Division of Molecular Medicine, Bijenicka cesta 54, 10000 Zagreb, Croatia; Department of Neurology. Complejo Hospitalario de Navarra, IdiSNA (Navarra Institute for Health Research), Pamplona, Spain; Public Health Laboratory in Gipuzkoa, Biodonostia Health Research Institute, San Sebastián, Spain

## Abstract

**Objective:** The lack of knowledge about the onset and progression of Parkinson’s disease (PD) hampers its early diagnosis and treatment. Our aim was to determine the biochemical remodeling induced by PD in a really early and pre-symptomatic stage and unveiling early potential diagnostic biomarkers adopting a multiplatform (LC-MS, GC-MS, CE-MS) untargeted metabolomics approach.

**Methods:** 41,437 healthy volunteers from the European Prospective Study on Nutrition and Cancer (EPIC)-Spain cohort were followed for around 15 years to ascertain incident PD. For this study, baseline pre-clinical plasma samples of 39 randomly selected individuals (46% females, 41– 69 years old) that developed PD (Pre-PD group) and the corresponding control group (*n*=39, 46% females, 41–69 years old) were analyzed. The metabolic differences were investigated by univariate and multivariate data analyses, followed by pathway-based metabolite analyses to obtain possible clues on biological functions.

**Results:** Our results exposed significantly lower levels of seven free fatty acids in the pre-PD subjects, together with alterations in other metabolite classes. Our finding revealed alterations in fatty acids metabolism, mitochondrial dysfunction, oxidative stress, and gut-brain axis dysregulation.

**Conclusions:** Although the biological purpose of these events is still unknown, the mechanisms involved in the remodelling of the suggested metabolic pathways seem to appear long before the development of PD hallmarks. These findings might be considered as worthy potential markers whose alteration might lead to the development of PD hallmarks in the future. Consequently, this study is of inestimable value since this is the first study conducted with samples collected many years before the disease development.

## 1. INTRODUCTION

Parkinson’s disease (PD) is a neurodegenerative disorder characterized by a loss of dopaminergic neurons in the substantia nigra and the production of Lewy bodies. Such neurological alterations cause motor and cognitive impairments^1 2^. Together with Alzheimer’s disease, it is one of the most prevalent neurodegenerative disorders with around 2% of affected people older than 60 years^3 4^. Diagnosis of PD is established according to the presence of parkinsonian motor symptoms, while therapy is exclusively symptomatic and cannot stop or slow down progression^5^. Since typical PD motor symptoms appear when there is already more than 80% of dopaminergic loss, identification of biomarkers for early diagnosis might drastically change diagnosis and treatment approaches^6^. Several approaches have already been proposed for the early diagnosis of PD, including positron emission tomography (PET) and single photon emission computed tomography (SPECT) imaging, examining olfactory alterations that usually appear before any known motor symptoms, skin and colonic biopsy, gene sequencing and changed metabolites, including uric acid, glutathione and α-synuclein^3^. Since these approaches fall short for early diagnosis due to the complexity of PD, novel diagnostic approaches that would combine several methods are necessary^3^. Moreover, a pre-symptomatic population has never been explored searching for biomarkers.

In this regard, metabolomics can be considered as a well-defined approach to unveil potential metabolic biomarkers to diagnose the disease when no PD symptoms have yet developed, to understand better its early pathophysiological mechanisms, to identify potential novel drugs, and to monitor the therapeutic outcome^7^. Numerous original metabolomics studies have been conducted using different biological samples in order to discover potential biomarkers and altered metabolic pathways in PD. Using different analytical techniques, alterations in amino acids, dysregulation of the TCA cycle, altered fatty acid, purine, and dopamine metabolism were observed in early, mid and advanced stages of PD^8 9^. Recently, an LC-MS metabolomics-based study conducted with plasma samples from PD patients reflected alterations in many metabolite classes, including a remarkable reduction of the levels of seventeen free fatty acids, cis-aconitic acid, and an increment on bile acids (BA) levels in PD patients^10^.

Here, we used mass spectrometry (MS) coupled to gas chromatography (GC), liquid chromatography (LC), and capillary electrophoresis (CE) to obtain a metabolome coverage as extensive as possible, aiming the analysis of the global metabolic changes in the plasma samples from participants from the European Prospective Study on Nutrition and Cancer (EPIC) that were followed for almost 15 years until June 2011. None of the participants had developed PD or showed any related symptoms at the time of sample collection. During the time they were followed up, some of them developed PD (pre-PD) while others did not (controls) (Figure 1). Consequently, these samples are of extraordinary value for discovering potential biomarkers for the early diagnosis of PD, conferring this metabolomics-based study a unique advantage in the field.

**Figure 1.**
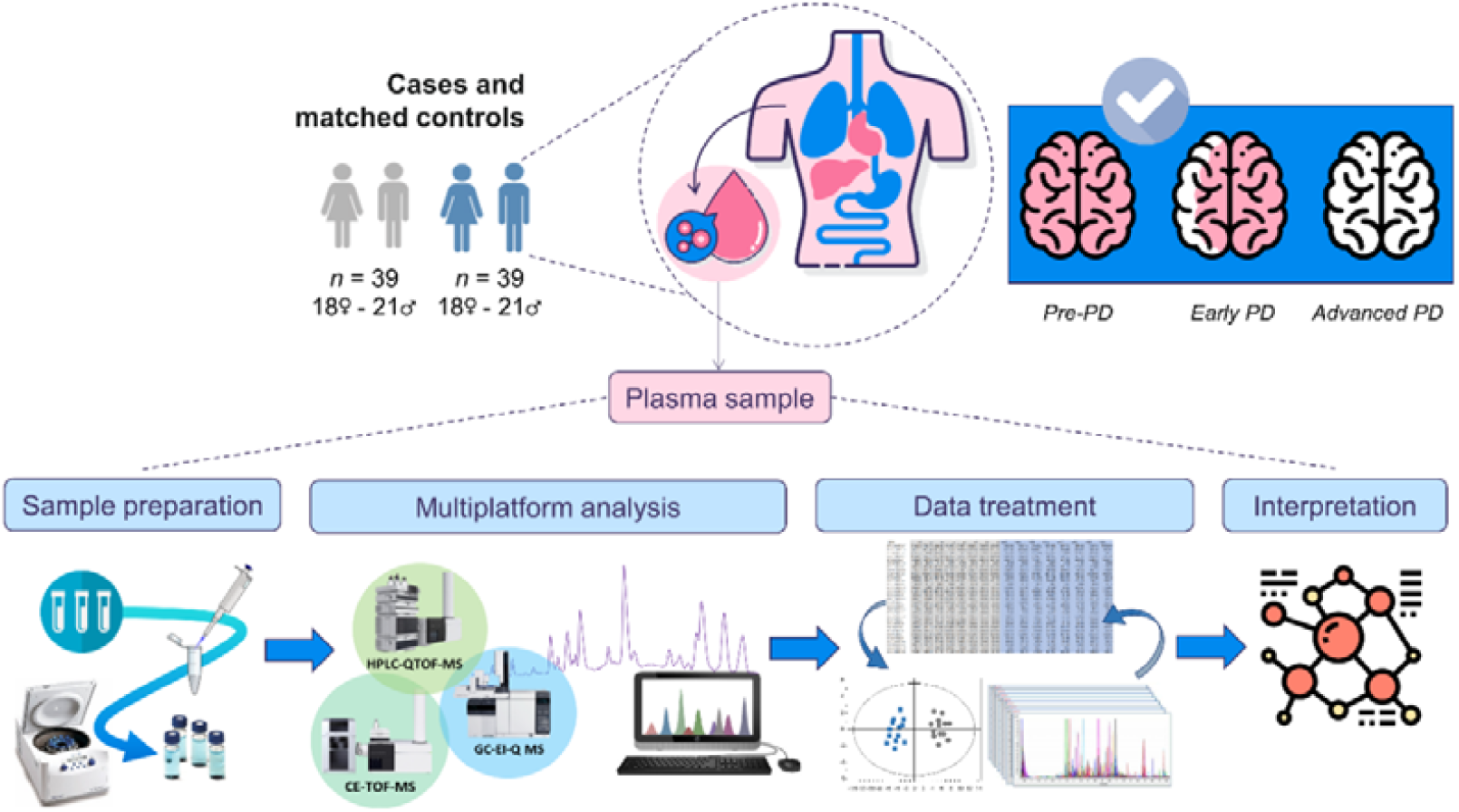
Metabolomics workflow followed for this study. Experimental design and the multiplatform untargeted metabolomics workflow followed for this study.

## 2. MATERIALS AND METHODS

### 2.1. Patients cohort and samples

The European Prospective Investigation into Cancer and Nutrition is an ongoing multicenter prospective cohort study designed to investigate the relationship between diet, nutrition and metabolic factors with cancer. Descriptions of study design, population and baseline data collection of the cohort have been reported in detail previously^11^. The Spanish EPIC cohort consists of 41,437 healthy participants (62% women), aged 29–69 years enrolled in 1993-1996. The EPIC-Spain Parkinson Cohort study comprises the three centres with available on Parkinson incident data Gipuzkoa, Navarra, and Murcia. The study sample consisted of 25,015 participants (57% women), aged 30–70 years at recruitment. Detailed description of lifestyle, diet, anthropometry and clinical data, case ascertainment, and the design of nested case-control study is collected in Supplementary Materials. For this multiplatform untargeted metabolomics-based study, 39 individuals that developed PD (Pre-PD group) and the corresponding control group were randomly selected, consisted both groups of 46% women, aged between 41 to 69 years old, and 54% males aged between 41 to 69 years old.

On the day of the interview subjects were appointed for blood sample analysis within the next week at the primary care centre, where the interview took place and a 30 ml blood sample were obtained at baseline. Fasting venous blood samples were drawn and immediately processed and were divided into 0.5 ml aliquots of plasma, serum, concentrated red blood cells and buffy coat, and stored in liquid nitrogen tanks at –190ºC until analysis. Quality assurance and plasma metabolites extraction was carried out according to previously reported standard protocols^12-15^ (Supplementary Materials).

### 2.2. Data treatment and statistical analyses

LC-MS and GC-MS raw data were processed using Agilent Technologies MassHunter Profinder B.08.00 SP1 software (Waldbronn, Germany) to obtain the data matrices. For GC-MS, deconvolution and identification were performed with Agilent MassHunter Unknowns Analysis Tool 9.0 using the Fiehn (v2013) and NIST (v2017) commercial libraries. The obtained GC-MS data was aligned using the MassProfiler Professional B.12.1 software (Agilent Technologies) and exported to Agilent MassHunter Quantitative Analysis version B.09.00 to assign the main ions and integrate each of the signals. Missing values were imputed using the K-nearest neighbors algorithm^16^. Afterward, the data matrices were filtered by coefficient of variation (CV), maintaining those signals that presented a CV < 30% in the QCs. The filtered matrices were imported into SIMCA P+15 (Umetrics, Umea, Sweden). The obtained PCA plot revealed an intra-batch effect due to gradual changes in the instrumental response that are often unavoidable, especially for long analytical sequences. In order to minimize the instrumental variation observed that hinders the power to detect the biological variation, the data were normalized by applying a correction called “quality control samples and support vector regression (QC-SVRC)”^17^. Furthermore, a supervised OPLS-DA model was obtained for the LC-MS ESI(+) data. The metabolites presenting a VIP ≥ 1 and jackknifing confidence interval not including the zero value were selected as statistically significant from the OPLS-DA model. Finally, OPLS-DA model was validated with cross-validation and CV-ANOVA tool provided by SIMPA-P+ software. The univariate data analysis was carried out using MATLAB R2015a software (Mathworks, Inc., Natick, USA), applying a t-test (p-value <0.05) and a standard Benjamini-Hochberg method was applied to control the false discovery rate (FDR) for multiple hypothesis testing. Finally, the percentage of change was calculated by comparing Pre-PD vs. control.

### 2.3. LC-MS and CE-MS metabolites annotation

The metabolites that turned out to be statistically significant (p-value <0.05) and showed % change greater than 20% were tentatively annotated. Initially, the m/z of the significant metabolites was searched against multiple databases available online, including METLIN (http://metlin.scripps.edu), LipidsMAPS (http://lipidMAPS.org) and KEGG (http://www.genome.jp/kegg/), through the CEU MassMediator (http://ceumass.eps.uspceu.es/)^18^. HMDB (http://hmdb.ca) was also consulted for additional information. Features that were tentatively assigned to metabolites from the databases were based on: (1) mass accuracy (maximum error mass 20 ppm), (2) isotopic pattern distribution, (3) possibility of cation and anion formation, (4) adduct formation, and (5) elution order of the compounds based on the chromatographic conditions. Additionally, an ‘in-house’ CE-MS library built with authentic standards was used to compare the relative migration time of the significant metabolites to increase the annotations confidence. As it can be observed in Table 1, five FFAs were detected as significantly affected by two independent analytical platforms (GC-MS and LC-MS) presenting the same trend. This fact ensures the great analytical performance and reproducibility of the analyses, increases the confidence level of the metabolite annotations, and verifies the consistency of the biological results throughout the whole study.

**Table 1.**
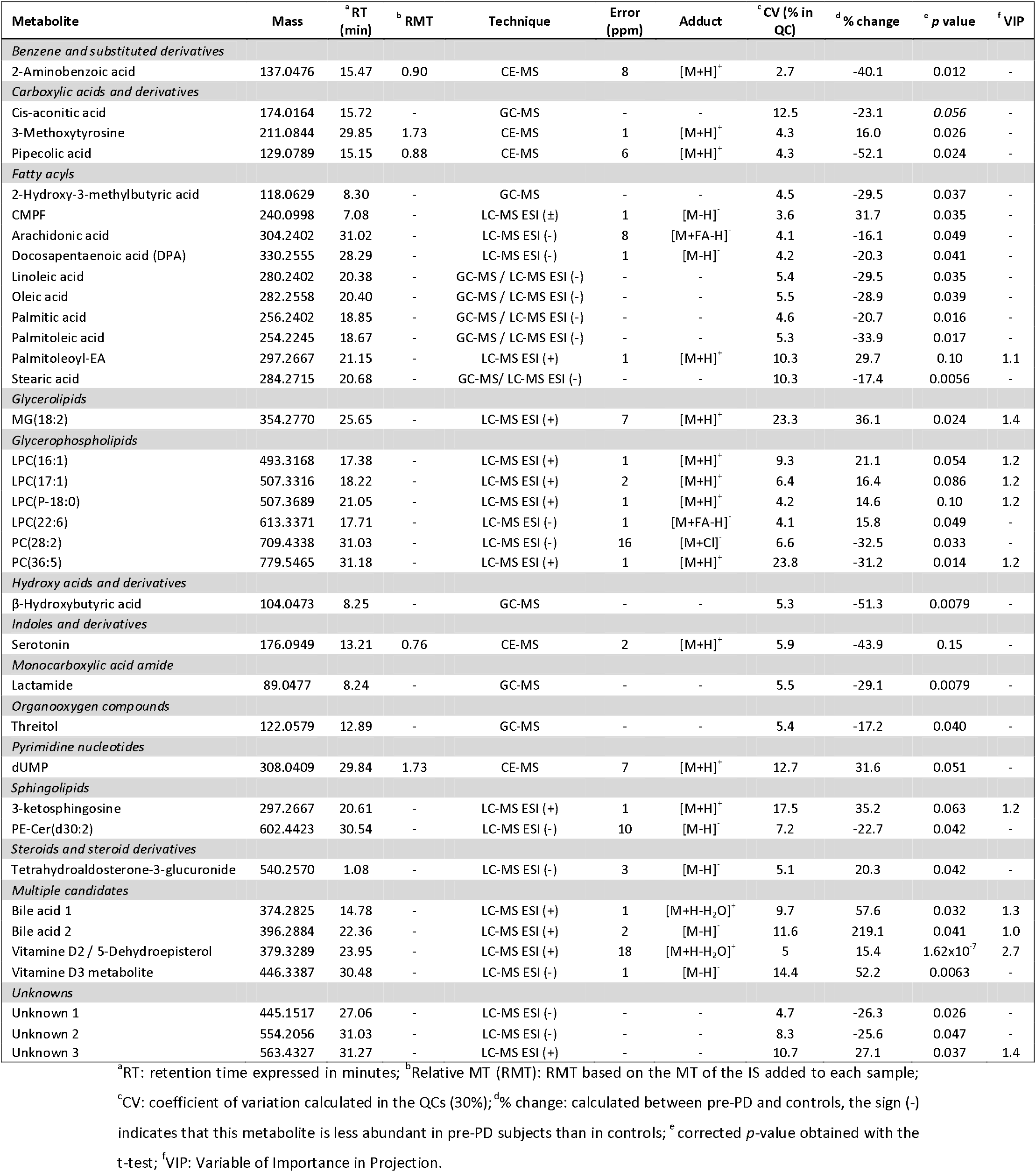
Metabolites that showed statistical significance when comparing PD subjects and healthy controls.

### 2.4. Identification of potential metabolite biomarkers

For biomarker prediction, the Multivariate ROC plot-based exploratory analysis (Explorer) was performed in MetaboAnalyst 4.0 (https://www.metaboanalyst.ca/) (Figure 2A-D). This analysis performs automated important feature identification and performance evaluation. The ROC curve analyses were based on Linear Support Vector Machine (SVM), and the ROC plots were generated by Monte-Carlo cross-validation using balanced sub-sampling. In each cross-validation, two thirds (2/3) of the samples are employed to evaluate the feature importance. The principal features are then exploited to construct the classification models, which are validated on the remaining 1/3 of the samples. The procedure is replicated multiple times to calculate the performance and the confidence interval of each model. For our data, the classification method selected was Linear SVM, and the feature ranking method selected was the SVM built-in algorithm^19^.

**Figure 2.**
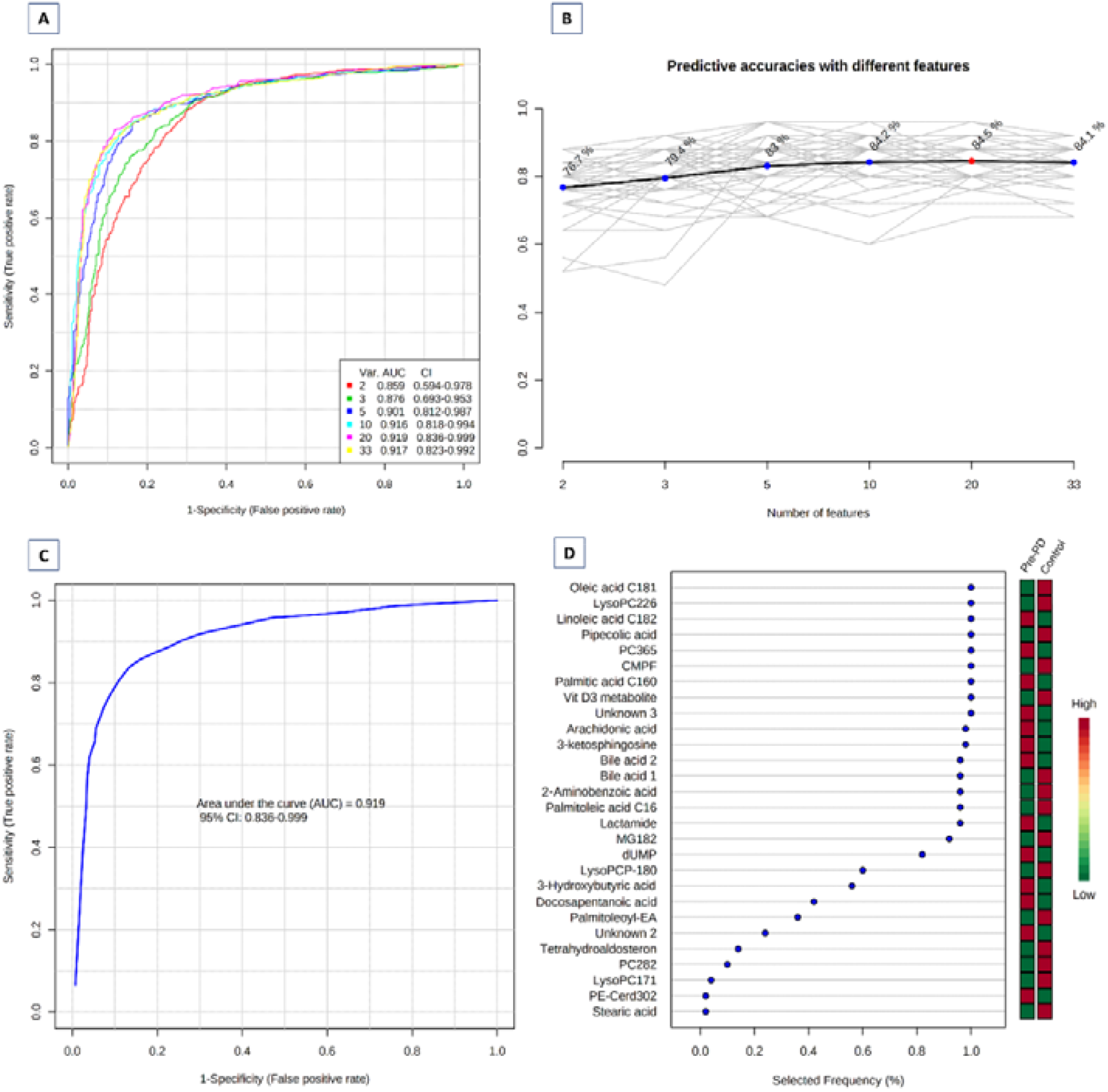
Biomarker prediction by Multivariate ROC curve based exploratory analysis. **A** Overview of all ROC curves created by MetaboAnalyst 4.0 from 6 different biomarker models considering different number of features (2, 3, 5, 10, 20, and 33) with their corresponding AUC value and confidence interval; **B** Predictive accuracies of 6 different biomarker models. The red dot specifies the highest accuracy for the 20-feature panel of model 5; **C** ROC curve for selected biomarker model 5; **D** Top 28 potential biomarkers predicted based on their frequencies of being selected during cross validation.

## 3. RESULTS

### 3.1. Metabolite coverage by a multiplatform untargeted metabolomics-based approach

The metabolic fingerprint of plasma samples collected several years before the development of PD symptoms was achieved by performing a multiplatform untargeted metabolomics strategy. The metabolic alterations unveiled in this study indicate that the pathological mechanisms behind the onset and progression of PD starts many years before the appearance of the first PD symptoms. A significant amount of information was obtained from the multiplatform analysis, resulting in 672 and 571 metabolic features detected in LC-MS operated in positive and negative ionization modes, respectively, 332 signals were acquired from CE-MS and 104 from GC-MS analysis. After raw data matrix normalization, curation, and statistical analysis, 15, 12, 9, and 3 metabolites were detected as significantly affected by LC-MS-ESI(-), LC-MS-ESI(+), GC-MS, and CE-MS, respectively (Table 1).

To enrich the biological interpretation of the results, we will discuss about the important changes displayed by cis-aconitic acid, 2’-deoxyuridine-5’-monophosphate (dUMP), and serotonin although they were not statistically significant. After data normalization, all the PCA plots obtained displayed a tightly clustering of the QC samples, revealing the correction of the instrumental variation (Figure S1). Surprisingly, a supervised OPLS-DA model was obtained for the LC-MS-ESI(+) data with great quality parameters values (R^2^ = 0.996, Q^2^ = 0.687)^20^. Finally, the models were validated by cross-validation and CV-ANOVA tool provided by SIMCA-P+ software (*p* CV-ANOVA=1.31×10^−8^) (Figure S2).

### 3.2. Metabolic alterations detected in plasma samples of pre-PD subjects

Most of the identified signals in LC-MS-ESI(+) were glycerophospholipids, while in LC-MS-ESI(-) most common metabolites were saturated (SFA), monounsaturated (MFA), and polyunsaturated (PUFA) fatty acids. The levels of four LPC species were increased, while PC(36:5) and PC(28:2) were significantly decreased in pre-PD group. Additionally, the BA levels were upregulated in pre-PD samples (Figure 3). However, the type of BA was not established according to the spectra and it was only identified according to the compound group. The Palmitoleoyl ethanolamide, MG(18:2), 3-ketosphingosine, and vitamin D2/5-dehydroepisterol were also increased in Pre-PD plasma samples. Regarding the LC-MS-ESI(-), the DPA, arachidonic, linoleic, oleic, stearic, palmitic, and palmitoleic acids were decreased in plasma samples of pre-PD subjects compared with controls, same as levels of tetrahydroaldosterone-3-glucuronide and CMPF. As in LC-MS-ESI(-), most of the significantly affected compounds obtained by GC-MS were also FFAs. Palmitoleic, linoleic, oleic, palmitic, and stearic acids were significantly decreased in Pre-PD plasma samples (Table 1) (Figure 3). Furthermore, β-hydroxybutyric acid, 2-hydroxy-3-methylbutyric acid, lactamide, and threitol levels were reduced in pre-PD subjects. Regarding CE-MS, the pipecolic acid and 2-aminobenzoic acid levels were decreased, while the 3-methoxytyrosin showed an increment in the plasma samples of pre-PD group. Finally, the potential biomarkers obtained from the multivariate ROC curve exploration module, oleic acid, LPC(22:6), linoleic acid, pipecolic acid, PC(36:5), CMPF, palmitic acid, vitamin D3 metabolite, arachidonic acid, 3-ketosphingosine, BA, 2-aminobenzoic acid, palmitoleic acid, lactamide, MG(18:2), and dUMP, presented a selection frequency between 0.8 and 1.0 (*i*.*e*. selected between 80% to 100% of the time in the model) of the SVM feature selection algorithm (Figure 2D).

**Figure 3.**
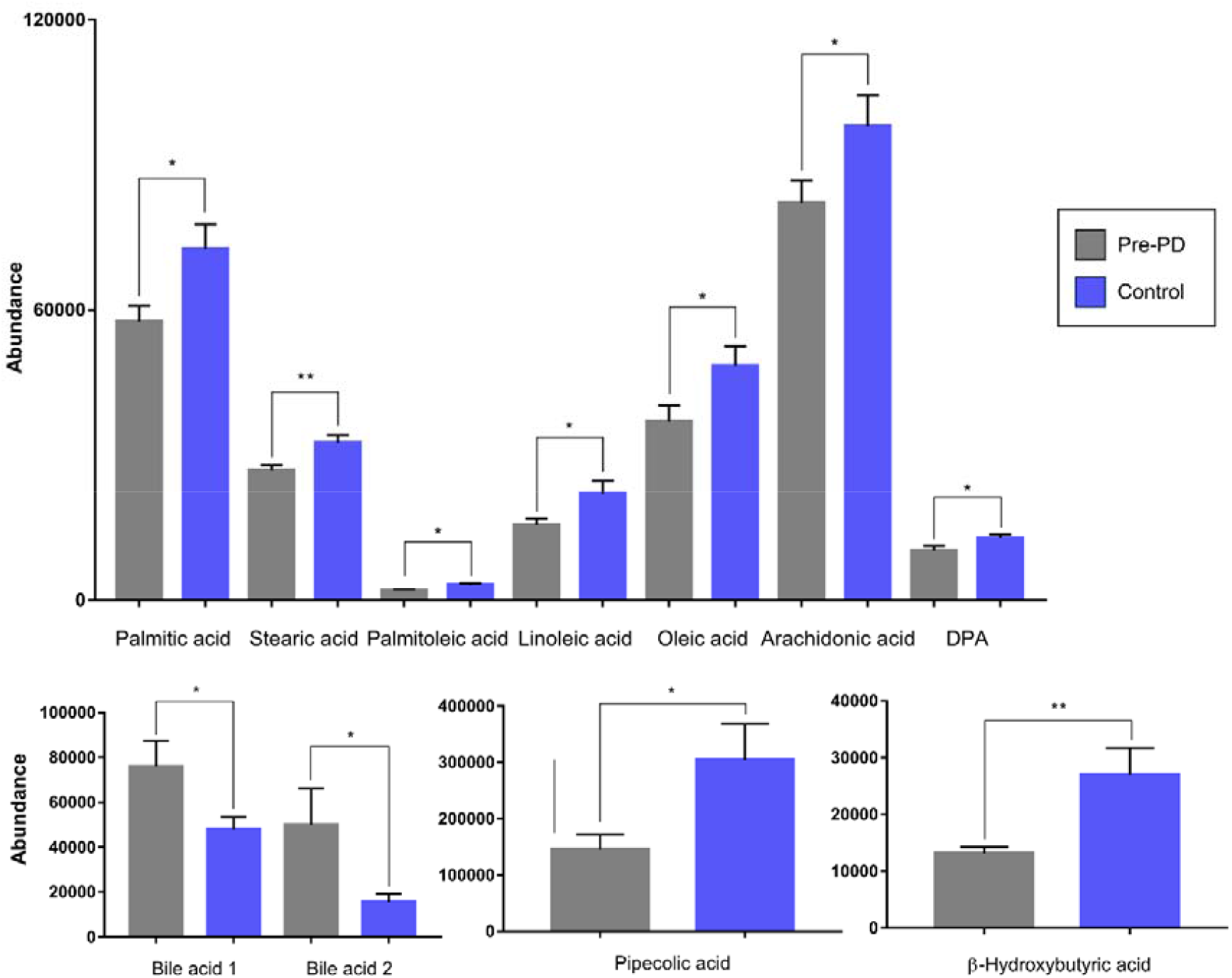
Abundance of the statistically significant FFAs, BA, pipecolic acid, and β-hydroxybutyric acid. The graphic reflects the differences between the pre-PD (grey bar) and the control (blue bar) group. The error bars represent the standard error of the mean (SEM). *p ≤ 0.05; **p ≤ 0.001.

## 4. DISCUSSION

Parkinson’s disease is a progressive, complex, multisystem neurodegenerative disorder, characterized by typical movement symptoms^21^ and cognitive impairments that affect usual functioning. Due to complexity and heterogeneity, several metabolic pathways are involved in its development and pathogenesis^9^. It is crucial to enlighten that metabolites mentioned below were already altered before PD development because the sampling was performed while subjects still did not have any symptoms.

### 4.1.1. Fatty acid metabolism alterations

In this study, decreased levels of multiple fatty acyls including SFA, MUFA, and PUFA were noticed in pre-symptomatic PD subjects (Figure 4). This is in agreement with recent studies that recognized a remarkable reduction of the levels of several FFAs in PD patients^8 10 22^.

**Figure 4.**
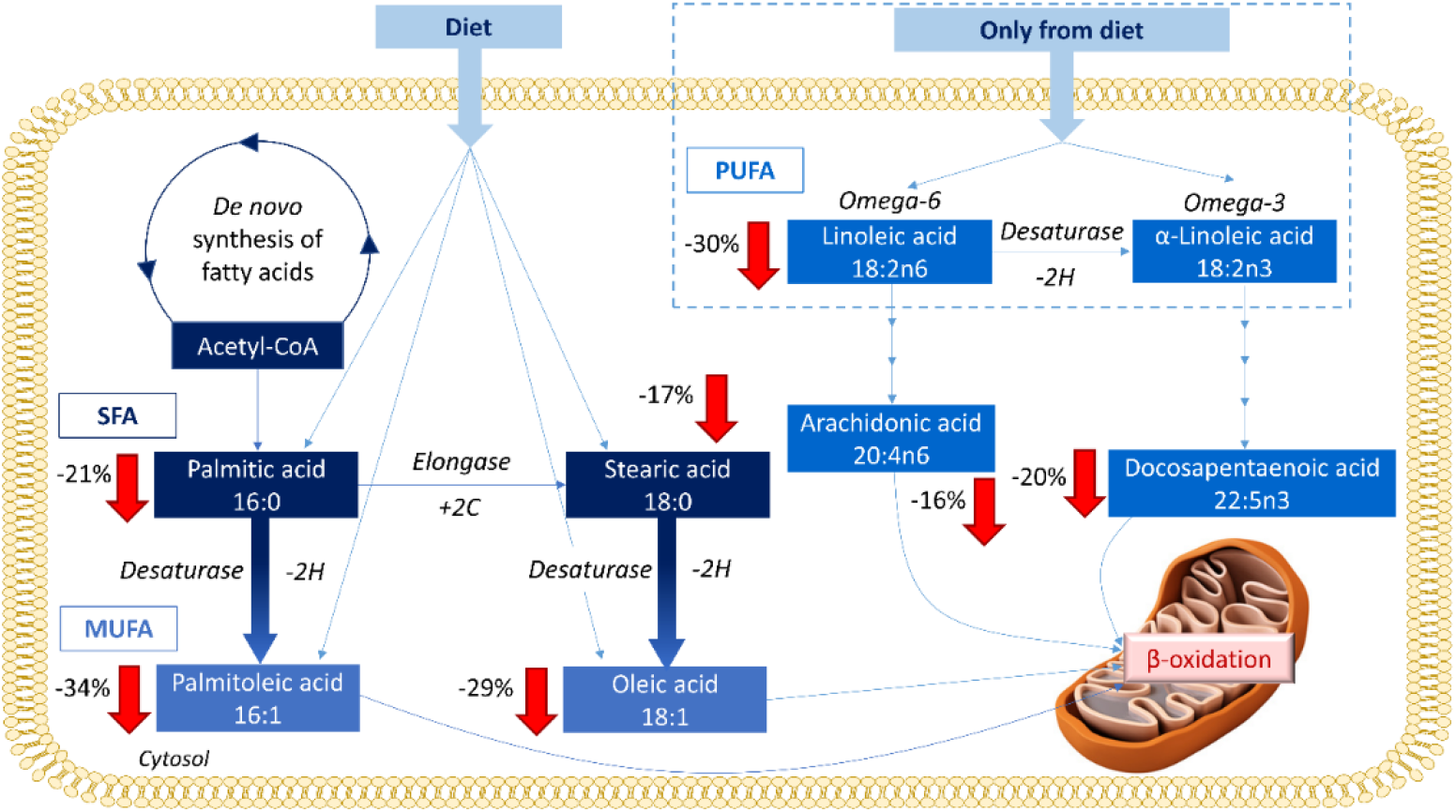
FFAs metabolism and their trends observed in this study when compared the pre-PD group with control subjects. The arrows represent the trend of each metabolite within the comparison, displaying their corresponding percentages of change.

Fatty acids are vital for a correct brain function since it depends on dietary fatty acid intake. Lipids and fatty acid dyshomeostasis is associated with many neurodegenerative disorders, neuroinflammation and oxidative stress^23^, same as with apoptotic signaling and mitochondrial dysfunction^8 9^. Currently, it seems that there is not direct connection between the SFA intake and PD risk in humans^24^. Higher palmitic and stearic acids levels have been observed in lipid rafts from the frontal cortex of PD patients compared to healthy controls^25^. Moreover, the α-synuclein modulates the uptake of palmitic acid into the brain^26^. Therefore, the accumulation of this protein in PD brains might lead to increased levels of palmitic acid, which in turn can trigger some of its neuropathological activities. Consequently, it can be hypothesized that the lower levels of both SFA in plasma of the pre-PD subjects might be due to an early and progressive migration and reuptake of these fatty acids to the brain. Regarding the MUFA, palmitoleic and oleic acids are recognized of great importance for human metabolism since they are considered anti-inflammatory compounds with neuronal protective effects^27^. Palmitoleic acid is one of the principal components of the human adipose tissue, muscle, and liver together with palmitic, stearic, oleic, and linoleic acids, whose levels have been also detected as significantly reduced in the PD group. A recent study has demonstrated that oleic acid presents detrimental effects in the α-synuclein homeostasis since it induces α-synuclein inclusion formation in human neural cells^28^. Therefore, the lower levels of oleic acid in plasma might be also due to its early reuptake by the brain. Regarding the PUFAs, although the epidemiological evidence suggesting that dietary fat consumption may be associated with PD risk is not consistent, a significant number of PD animal models and epidemiological research studies have proved that PUFAs might play many critical roles in this regard^29^. The elevated PD risk may result from the dietary fat effects, including increased oxidative stress and neuroinflammation, which can potentially worsen the dopaminergic neuron loss due to neurotoxicity^29^.

Although increased levels of SFAs has been observed in a 6-OHDA rat model of PD^30^, a recent study conducted with plasma samples collected from PD patients’ uncovered diminished levels of 4 SFAs, including the stearic acid in the PD group compared with their corresponding control group^10^. The study also revealed reduced levels of 4 MFA, including the oleic acid, and 9 PUFA, including the linoleic acid, arachidonic acid, and DPA^10^.

### 4.1.2. Pipecolic acid and the gut-brain axis

It is becoming increasingly evident that intestinal microbiota influences gut-brain axis communication. Indeed, the premise that PD development is triggered following continuous gut aggravation has gathered significant strength recently^31^. In fact, the enteric αSyn has been associated with greater intestinal permeability^32^. It exists a positive relationship between inflammatory bowel diseases and future PD risk in various populations^33^. A previous study demonstrated how metabolites involved in the correct brain function were influenced by the intestinal microbiota, including the pipecolic acid^34^. This metabolite comes from the degradation of lysine in the cerebral peroxisomes. These multifunctional organelles are involved in ROS metabolism, fatty acid oxidation, ether lipid synthesis, BA synthesis, and cholesterol transport. Peroxisomal dysfunction has been linked to neurodegenerative disorders, including Parkinson’s disease (PD)^35^. Pipecolic acid can act also as neurotransmitter modulating the gamma-aminobutyric acid (GABA)ergic transmission^36^. It can be taken up by cerebral mitochondria, the principal responsible of cellular apoptosis activation, stimulating the neuronal cell death^36^. Therefore, decreased levels of pipecolic acid might be indicative of the initial stages of the gut-brain axis dysregulation and a peroxisomal failure. Additionally to pipecolic acid, we did not observe any other significantly changed amino acid, while other articles reported alterations on their levels in PD subjects^8 9^. As our study samples were collected prior to disease development, changes in these compounds might be considered as metabolic biomarkers of advanced stages of the PD.

### 4.1.3. Ketone bodies and mitochondrial function impairment

We found decreased β-Hydroxybutyric acid levels in Pre-PD cohort. This ketone body is synthesized in liver mitochondria from acetyl-CoA and serves as an alternative fuel source for extrahepatic tissues including the brain. It is a product of healthy metabolism of fatty acid oxidation together with acetoacetate. It has been described that β-Hydroxybutyric acid administration protects both mesencephalic and dopaminergic neurons from MPP^+^ and MPTP toxicity, respectively^37^. Moreover, patients able to tolerate a ketogenic diet improved their Unified Parkinson Disease Rating Scale scores with no harmful effects^38^. Consequently, a decrease in this metabolites levels might indicate the initial stage of the mitochondrial impairment that plays a central role in the PD pathophysiology.

### 4.1.4. Energy production alterations

Increased levels of BA have also been observed. In accordance with our results, it has been recently described an increment of BA levels in plasma samples of PD patients^10^. Consequently, elevations of BA levels can also be detected many years before the PD development. Due to their role, it is assumed that down-regulation of fatty acids and phospholipids and other perturbations in lipid metabolism might be associated with impaired BA metabolism. Consequently, it could cause further disturbances in energy production in PD subjects^39^. Together with fatty acid metabolism, TCA cycle plays an important role in energy metabolism. Dysregulation of TCA cycle is possibly associated with PD progression and α-synuclein pathology, while fatty acid metabolism might be associated with α-synuclein aggregation^9^. We identified decreased levels of cis-aconitic acid in Pre-PD subjects. It is assumed that TCA cycle has an important role as part of energy metabolism in degeneration of dopaminergic cells through complex *gene x environment* interaction^9^. At the beginning of neurodegenerative processes in PD, TCA cycle is downregulated due to mitochondrial dysfunction or energy deficient^9^, which is in correspondence with downregulation of fatty acid metabolites also due to disturbances in mitochondrial function and energy production^39^. Although the difference between pre-PD subjects and controls was not significant in our study, it has been newly uncovered diminished cis-aconitic acid levels in PD patients^10^. Therefore, reduced levels of TCA cycle metabolites might be related to the future onset of PD.

### 4.1.5. Tryptophan metabolism and PD onset

The metabolism of tryptophan is associated with PD progression and development^9^. Postmortem examinations in PD patients showed decreased levels of metabolites involved in tryptophan pathway, same as altered tryptophan levels in CSF and plasma. It is assumed that dysregulation of this metabolism leads to neurotoxicity^40^, which might act as a trigger for PD development. This study has shown decreased levels of tryptophan co-metabolites, such as 2-aminobenzoic acid and serotonin in PD patients. Unlike serotonin, 2-aminobenzoic acid was significantly decreased in PD subjects, which might confirm possible association of altered tryptophan metabolism in early stage of disease development.

## 5. CONCLUSIONS

Through a multiplatform untargeted metabolomics-based approach we have identified, for the first time, 33 altered metabolites in plasma samples from subjects that did not present any pathology when the samples were collected and, many years later, they developed PD. Several pieces of evidence indicate the implication of mitochondrial dysfunction and oxidative stress, which is reflected in the metabolite changes of PD pathogenesis. According to our data, it is evident that changes in fatty acid metabolism and their corresponding metabolic pathways are altered long before PD’s first symptoms are observed. Likewise, in our study, these metabolites levels were significantly reduced compared to subjects that did not develop PD during the time they were followed up. Our results are mostly in agreement with recent metabolomics-based findings where decreased levels of 17 FFAs and cis-aconitic acid and increased BA levels were found in plasma samples collected from PD patients. This fact indicates that these metabolite modifications begin many years before the PD development and are maintained when suffering this disease. Furthermore, the detected alteration in pipecolic acid levels leads to hypothesize about the early triggering of the gut-brain axis dysregulation and peroxisomal failure. Although the biological purpose of these events is still unknown, the mechanisms involved in the suggested mitochondrial dysfunction, oxidative stress, and gut-brain axis dysregulation seem to appear long before the development of this disease. Therefore, the remodeled metabolic pathways highlighted in this study might be considered as worthy potential markers whose alteration might lead to the development of PD hallmarks in the future.

## Data Availability

The datasets used and/or analyzed during the current study are available from the corresponding author on reasonable request.

## Abbreviations

BA: bile acids
CMPF: 3-carboxy-4-methyl-5-propyl-2-furanpropionic acid
DPA: docosapentaenoic acid
EPIC: European Prospective Study on Nutrition and Cancer
FFAs: free fatty acids
LPC: lysoglycerophosphocholines
MFA: monounsaturated fatty acid
MPP^+^: 1-Methyl-4-phenylpyridinium
MPTP: 1-methyl-4-phenyl-1,2,3,6-tetrahydropyridine
PC: glycerophosphocholines
pre-PD: pre-Parkinson’s Disease development group
PUFA: polyunsaturated fatty acid
QC-SVRC: quality control samples and support vector regression
RMT: relative migration time
SFA: saturated fatty acid
SVM: Support Vector Machine.

## DECLARATIONS

### Ethics approval and consent to participate

This study was approved by the Ethics Committee for clinical research of the Basque Country PI2017031.

### Authors’ Contributions

AB, PA, JMH, EA, FGI, CB were involved in study concept and design. JS, MK, CGR, CB, JMH, EA, PA, AB, MDC were involved in data acquisition. CGR and CB were involved in data analysis. CGR, JS, CB, AB, PA, JMH, EA, FGI, EA were involved in data interpretation. CB, AB, FGI, PA were involved in the work supervision. CGR, JS, MK, CB were involved in drafting the manuscript. CGR, CB, MEE, EM, AB, PA, JMH, EA, FGI, EA, MDC were involved in critical revision of the manuscript for important intellectual content. All authors read and approved the final manuscript.

## SUPPLEMENTARY MATERIALS

### Additional file

- Study population details – page 4
- Chemicals, metabolites extraction protocols, and quality assurance – pages 7-8
- Run settings and data acquisition for LC-MS, GC-MS, and CE-MS – page 9
- Strengths and limitations of the study – page 12
- Figure S1. PCA-X score plot for the 3 analytical platforms – page 13
- Figure S2. Supervised OPLS-DA plot of LC-MS (+) data analysis – page 13

## REFERENCES

1. Han W, Sapkota S, Camicioli R, et al. Profiling novel metabolic biomarkers for Parkinson’s disease using in-depth metabolomic analysis. Movement Disorders 2017;32(12):1720–28.

2. Sveinbjornsdottir S. The clinical symptoms of Parkinson’s disease. Journal of neurochemistry 2016;139:318–24.

3. De Virgilio A, Greco A, Fabbrini G, et al. Parkinson’s disease: autoimmunity and neuroinflammation. Autoimmunity reviews 2016;15(10):1005–11.

4. Bose A, Beal MF. Mitochondrial dysfunction in Parkinson’s disease. Journal of neurochemistry 2016;139:216–31.

5. Cookson MR. Chapter 6 - Parkinson’s disease. In: Baekelandt V, Lobbestael E, eds. Disease-Modifying Targets in Neurodegenerative Disorders: Academic Press 2017:157–74.

6. Cheng HC, Ulane CM, Burke RE. Clinical progression in Parkinson disease and the neurobiology of axons. Annals of neurology 2010;67(6):715–25.

7. Wishart DS. Emerging applications of metabolomics in drug discovery and precision medicine. Nature Reviews Drug Discovery 2016;15(7):473–84. doi: 10.1038/nrd.2016.32

8. Havelund JF, Heegaard NH, Færgeman NJ, et al. Biomarker research in Parkinson’s disease using metabolite profiling. Metabolites 2017;7(3):42.

9. Shao Y, Le W. Recent advances and perspectives of metabolomics-based investigations in Parkinson’s disease. Molecular neurodegeneration 2019;14(1):3.

10. Shao Y, Li T, Liu Z, et al. Comprehensive metabolic profiling of Parkinson’s disease by liquid chromatography-mass spectrometry. Molecular neurodegeneration 2021;16(1):1–15.

11. Riboli E, Hunt K, Slimani N, et al. European Prospective Investigation into Cancer and Nutrition (EPIC): study populations and data collection. Public health nutrition 2002;5(6b):1113–24.

12. Naz S,Calderón ÁA, García A, et al. Unveiling differences between patients with acute coronary syndrome with and without ST elevation through fingerprinting with CE-MS and HILIC-MS targeted analysis. Electrophoresis 2015;36(18):2303–13.

13. Garcia A, Barbas C. Gas chromatography-mass spectrometry (GC-MS)-based metabolomics. Metabolic Profiling: Springer 2011:191–204.

14. Ciborowski M, Javier Ruperez F, Marnez-Alcázar MP, et al. Metabolomic approach with LC− MS reveals significant effect of pressure on diver’s plasma. Journal of Proteome Research 2010;9(8):4131–37.

15. Dudzik D, Barbas-Bernardos C, García A, et al. Quality assurance procedures for mass spectrometry untargeted metabolomics. a review. Journal of pharmaceutical and biomedical analysis 2017

16. Armitage EG, Godzien J, Alonso-Herranz V, et al. Missing value imputation strategies for metabolomics data. Electrophoresis 2015;36(24):3050–60.

17. Kuligowski J, Sánchez-Illana Á, Sanjuán-Herráez D, et al. Intra-batch effect correction in liquid chromatography-mass spectrometry using quality control samples and support vector regression (QC-SVRC). Analyst 2015;140(22):7810–17.

18. Gil-De-La-Fuente A, Godzien J, Saugar S, et al. CEU Mass Mediator 3.0: A Metabolite Annotation Tool. Journal of Proteome Research 2019;18(2):797–802.

19. Barberini L, Noto A, Saba L, et al. Multivariate data validation for investigating primary HCMV infection in pregnancy. Data in brief 2016;9:220–30.

20. Godzien J, Ciborowski M, Angulo S, et al. From numbers to a biological sense: How the strategy chosen for metabolomics data treatment may affect final results. A practical example based on urine fingerprints obtained by LC-MS. Electrophoresis 2013;34(19):2812–26. doi: 10.1002/elps.201300053

21. Emamzadeh FN, Surguchov A. Parkinson’s disease: biomarkers, treatment, and risk factors. Frontiers in neuroscience 2018;12:612.

22. Schmid SP, Schleicher ED, Cegan A, et al. Cerebrospinal fluid fatty acids in glucocerebrosidase-associated Parkinson’s disease. Movement disorders 2012;27(2):288–93.

23. Willkommen D, Lucio M, Moritz F, et al. Metabolomic investigations in cerebrospinal fluid of Parkinson’s disease. PloS one 2018;13(12)

24. Miyake Y, Sasaki S, Tanaka K, et al. Dietary fat intake and risk of Parkinson’s disease: a case-control study in Japan. Journal of the neurological sciences 2010;288(1-2):117–22.

25. Fabelo N, Martín V, Santpere Baró G, et al. Severe alterations in lipid composition of frontal cortex lipid rafts from parkinson’s disease and incidental parkinson’s. Molecular Medicine, 2010, vol 17, num 9-10, p 1107–1118 2010

26. Golovko MY, Faergeman NJ, Cole NB, et al. α-synuclein gene deletion decreases brain palmitate uptake and alters the palmitate metabolism in the absence of α-synuclein palmitate binding. Biochemistry 2005;44(23):8251–59.

27. Frigolet ME, Gutiérrez-Aguilar R. The role of the novel lipokine palmitoleic acid in health and disease. Advances in Nutrition 2017;8(1):173S–81S.

28. Fanning S, Haque A, Imberdis T, et al. Lipidomic analysis of α-synuclein neurotoxicity identifies stearoyl CoA desaturase as a target for Parkinson treatment. Molecular cell 2019;73(5):1001-14. e8.

29. Qu Y, Chen X, Xu M-M, et al. Relationship between high dietary fat intake and Parkinson’s disease risk: a meta-analysis. Neural regeneration research 2019;14(12):2156.

30. Shah A, Han P, Wong M-Y, et al. Palmitate and Stearate are Increased in the Plasma in a 6-OHDA Model of Parkinson’s Disease. Metabolites 2019;9(2):31.

31. Foster JA, Neufeld K-AM. Gut–brain axis: how the microbiome influences anxiety and depression. Trends in neurosciences 2013;36(5):305–12.

32. Forsyth CB, Shannon KM, Kordower JH, et al. Increased intestinal permeability correlates with sigmoid mucosa alpha-synuclein staining and endotoxin exposure markers in early Parkinson’s disease. PloS one 2011;6(12)

33. Gorecki AM, Preskey L, Bakeberg MC, et al. Altered gut microbiome in Parkinson’s disease and the influence of lipopolysaccharide in a human α-synuclein over-expressing mouse model. Frontiers in neuroscience 2019;13:839.

34. Matsumoto M, Kibe R, Ooga T, et al. Cerebral low-molecular metabolites influenced by intestinal microbiota: a pilot study. Frontiers in systems neuroscience 2013;7:9.

35. Cipolla CM, Lodhi IJ. Peroxisomal dysfunction in age-related diseases. Trends in Endocrinology & Metabolism 2017;28(4):297–308.

36. Matsumoto S, Yamamoto S, Sai K, et al. Pipecolic acid induces apoptosis in neuronal cells. Brain research 2003;980(2):179–84.

37. Tieu K, Perier C, Caspersen C, et al. D-β-Hydroxybutyrate rescues mitochondrial respiration and mitigates features of Parkinson disease. The Journal of clinical investigation 2003;112(6):892–901.

38. VanItallie TB, Nonas C, Di Rocco A, et al. Treatment of Parkinson disease with diet-induced hyperketonemia: a feasibility study. Neurology 2005;64(4):728–30.

39. Zhao H, Wang C, Zhao N, et al. Potential biomarkers of Parkinson’s disease revealed by plasma metabolic profiling. Journal of Chromatography B 2018;1081:101–08.

40. Szabó N, Kincses ZT, Toldi J, et al. Altered tryptophan metabolism in Parkinson’s disease: a possible novel therapeutic approach. Journal of the neurological sciences 2011;310(1-2):256–60.

