## Supplementary Materials for "Searching for prognostic biomarkers of Parkinson’s Disease development in the Spanish EPIC cohort through a multiplatform metabolomics approach"

**CONTENTS**


- Study population details – page 4
- Chemicals, metabolites extraction protocols, and quality assurance – pages 7-8
- Run settings and data acquisition for LC-MS, GC-MS, and CE-MS – page 9
- Strengths and limitations of the study – page 12
- Figure S1. PCA-X score plot for the 3 analytical platforms – page 13
- Figure S2. Supervised OPLS-DA plot of LC-MS (+) data analysis – page 13

**Study population**

The Spanish EPIC cohort consists of 41,437 participants (62% women), aged 29–69 years, enrolled in five Spanish regions: three in the North (Asturias, Gipuzkoa and Navarra) and two in the South (Granada and Murcia) between 1993-1996. The participants were healthy volunteers, mostly blood donors (67%) but also employees from private companies (3%), civil servants (5%), and the general population (23%) all of whom were fully covered by the public health system. The exclusion criteria were pregnancy, lactation and not being physically or mentally capable of participating [1].

*1.1. Lifestyle, Diet, Anthropometry and Clinical Data*

Baseline dietary and lifestyle data was collected in face-to-face interviews using validated questionnaires. Detailed descriptions of both the dietary and lifestyle questionnaires used have been published previously. A lifestyle questionnaire was used to collect information on sociodemographic characteristics, lifestyle and medical history as well as on reproductive factors in women. Anthropometric measures, height, weight, and waist circumference were measured in all of the participants using standard procedures. Body mass index was computed as weight (in kilograms) divided by height (in meters) squared [2, 3].

*1.2. Case Ascertainment*

Potential cases were identified using different sources of information depending on the centre. Each centre used at least two of the following: 1) record linkage with Primary Health records using whether the ICD-9 codes 332 or the International Classification of Primary Care (ICPC) codes N87 for Parkinson's disease; 2) record linkage with Prescriptions registry including subjects with at least one prescription of any of the N04-Antiparkinson Drugs of the ATC/DDD index (N04 - Antiparkinson Drugs; N04A - Anticholinergic agents; N04B - Dopaminergic agents); 3) record linkage with mortality registry using the ICD-9 codes 332 for PD; 4) hospital discharge database using the ICD-9 codes 332 for PD; 5) death certificates using the ICD-10 code G20.

Subjects identified in each of these sources were linked on EPIC identifier and duplicates discarded.

After a follow-up period running from recruitment to June 2011, about 90 individuals developed PD. Each diagnosis was based on a matrix combining two variables: the amount and quality of data available, and the degree of confidence of the neurologist expert in movement disorders reviewing the evidence. Diagnoses were defined as “definite” only when the degree of confidence of the neurologist was high and data quality excellent; diagnoses were defined as “very likely” in case the degree of confidence of the neurologist was high, but data quality was either good or poor; and defined as “probable” when the degree of confidence of the neurologist was medium and data quality was either excellent or good; finally, diagnoses were defined as “possible” in all remaining cases. The centers of Navarra and Murcia were able to verify all potential cases and San Sebastian could verify up to 84.8% of potential cases.

Vital status of the cohort and cause of death were assessed through record linkage with the regional mortality registry and the Spanish National Statistics Institute (www.ine.es).

### *1.3. Design of nested case-control study*

For each case subject up to two control subjects were randomly selected among appropriate risk sets consisting of all female cohort members with a blood sample, alive and free of cancer at the time of diagnosis of the index case. An incidence density sampling protocol was used, such that, in principle, control subjects could include study participants who became a case later in time and each control subject could be sampled more than once – the control subjects actually drawn, however, did not include any of the future cases of ovarian cancer detected so far in the EPIC cohort. Case and control subjects were matched on study recruitment centre, age at blood donation (±6 months), time of the day of blood collection (±1 h), fasting status (<3 h, 3–6 h, >6 h), follow-up time, and menopausal status at blood collection (premenopausal, perimenopausal, postmenopausal), current use of exogenous hormones (oral contraceptives, HRT) at the time of blood draw, as well as menstrual cycle phase for premenopausal women (3-5 categories, depending on available data). Cases missing data on phase of menstrual cycle were matched to control subjects whose information on menstrual cycle phase was also missing.

**Metabolites Extraction**

*Chemicals*

Organic solvents (MS grade), analytical grade formic acid 99%, standard mix for GC-MS containing grain fatty acid methyl ester (FAME) mixture (C8:0–C22:1n9), methanol, ethanol, and tricosane were from Sigma-Aldrich. Analytical grade heptane was purchased from Fluka Analytical (Sigma-AldrichChemie GmbH, Steinheim, Germany). Sialylation-grade pyridine was from VWR International BHD Prolabo (Madrid, Spain). Reference mass solutions for LC-MS and CE-MS were from Agilent Technologies. Ultrapure water (Milli-Qplus185 system Millipore, Billerica, MA, USA) was used in preparation of all buffers and standard solutions.

*Liquid chromatography coupled with mass spectrometry*

The 78 randomly selected plasma samples (pre-PD group, *n* = 39; control group, *n* = 39; gender balanced) were thawed on ice for approximately 1 hour. The samples were vortex-mixed for 2 minutes and 100 µL of plasma were transferred to an Eppendorf tube. Subsequently, 300 µL of a previously prepared cold mixture (-20°C) of methanol:ethanol (1:1, v/v) were added in the Eppendorf tube for deproteinization. After stirring the samples for 1 minute, they were incubated on ice for 5 minutes and vortex-mixed for another minute. Samples were centrifuged for 20 minutes at 13,000 rpm at 4 °C. After centrifugation, 100 µL of the supernatant was transferred to a chromatography vial with insert and was directly injected into the system.

*Gas chromatography coupled with mass spectrometry*

Once the samples were thawed on ice for 1 hour, they were vortex-mixed for 2 minutes and 40 µL of plasma were transferred to an Eppendorf tube. A volume of 120 µL of cold acetonitrile (-20 °C) was used for deproteinization. Sample preparation continued with vortex mixing for 2 minutes, incubation on ice and centrifugation for 10 minutes at 15,400 rpm at 4$^{o}$C. Then, 100 µL of the supernatant were transferred to a GC-MS vial to be evaporated to dryness in a vacuum concentrator. Once the vials were completely dry, the derivatization process was continued to obtain volatile derivatives for analysis. First, 10 µL of *O*-methoxyamine in pyridine (15mg/mL) were added to each of the vials. The samples were then vortex-mixed vigorously for 5 minutes, and then 3 sonication cycles (2 minutes) and 3 vortex cycles (2 minutes) were performed. The samples were covered with aluminum foil and incubated at room temperature, in the dark to complete the methoximation process. After 16 hours of incubation, 10 µL of N,O bis(trimethylsilyl)trifluoroacetamide (BSTFA) with 1% trimethylchlorosilane (TMCS) were added to all the samples, followed by 5 minutes of vortex-mixing. Samples were placed in the oven at 70 °C for 1 hour to complete the silylation reaction, which was followed by 30 minutes of cooling down. Finally, 100 µL of heptane with 20 ppm of tricosane (internal standard) were added to the samples and mixed for 1 minute in the vortex mixer.

*Capillary electrophoresis coupled with mass spectrometry*

After thawing the samples on ice for 1 hour, they were vortex-mixed for 1 minute and 100 µL of plasma were transferred to an Eppendorf tube. Subsequently, 100 µL of 0.2 M formic acid containing 5% acetonitrile and 0.4 mM methionine sulfone as an internal standard were added. The samples were vortex-mixed for 1 minute and filtered with a Millipore filter to remove proteins. Finally, the samples were centrifuged for 70 minutes, at 2000 rpm, at 4 °C. After centrifugation, 90 µL of the supernatant were transferred to a CE-MS vial for analysis.

*Quality control (QCs) samples and blanks*

Quality control samples (QC) were prepared by pooling and mixing equal volumes of each plasma sample to check the performance of the systems and the reproducibility of the sample treatment. Then, samples were randomized, and QCs were injected at the beginning, along the sequence, and at the end of the batch. Finally, two blank solutions were prepared along with the rest of the samples and analyzed at the beginning and at the end of the analytical sequence

**Analytical setup**

1. *Analytical settings for the LC-MS analysis*

The analysis of the samples was accomplished using an UHPLC system (1200 Infinity system, Agilent Technologies, Waldbronn, Germany), coupled to a 6520 QTOF MS (Agilent Technologies) with an ESI ion source. The sample injection volume was set up to 10 µL. The separation was achieved using a Discovery® HS C18 15cm x 2.1 mm, 3 µm (Supelco analytical) reverse phase column at thermostated 40 °C. The gradient used for the analysis consisted of a mobile phase A (0.1% formic acid in Milli-Q water) and a mobile phase B (0.1% formic acid in acetonitrile) pumped at 0.6 mL/min. The chromatography gradient began with 25% of phase B, increasing to 95% B in minute 35. The gradient then decreased to 25% of B in minute 36 and was maintained for 9 minutes until minute 45. Data were collected in positive and negative ESI modes in separate analyses, operated in full scan mode with a mass range of 50 to 1000 m/z for both modes. The capillary voltage was set to 3500 V for positive and 4000 V for negative ionization mode, the drying gas flow rate was 10.5 L/min at 330 ° C, gas nebulizer at 52 psi, and the fragment voltage 175 V. Two reference masses were used per ionization mode in order to provide a constant mass correction: m/z 121.0509 and m/z 922.0098 for the positive ionization mode, and m/z 119.0363 and m/z 966.0007 for the negative mode [4, 5].

2. *Analytical settings for the GC-MS analysis*

An Agilent GC system (7890A) coupled to a- 5975C mass spectrometer (Agilent Technologies) was used to perform metabolite fingerprinting of plasma samples. Briefly, 2 µL of derivatized samples were automatically injected in split mode (ratio 1:10) through a split liner of ultra-inert deactivated glass wool from Agilent. The separation of the compounds was achieved using a pre-column (10 m J&W integrated with Agilent 122-5532G) combined with a GC DB5-MS column (length, 30 m; internal diameter, 0.25 mm; and 0.25 µm film of 95% of dimethyl/5% diphenylpolysiloxane). The flow rate of the carrier gas (helium) was constant at 1 mL/min through the column. The retention time (RT) was locked according to the peak of the internal standard (C18 methyl stearate) at 19.66 minutes. The temperature of the column was initially set at 60 °C for 1 minute, then raised to 10 °C/min to 325 °C, which was maintained for 10 minutes before cooling. The injector and transfer line temperatures were set at 250 °C and 280 °C, respectively. The operating parameters of electronic impact ionization were established as follows: filament source temperature at 230 ° C and electronic ionization energy at 70 eV. Mass spectra were collected in a mass range of 50 to 600 m/z at a scan rate of 2 spectra per second. Data was acquired using Agilent MSD ChemStation software (Agilent Technologies). To determine the retention rate, a mixture of n-alkanes (C8-C28) dissolved in n-hexane was analyzed before the samples.

### 3. *Analytical settings for the CE-MS analysis*

The analysis was performed using a 7100 capillary electrophoresis (Agilent Technologies) coupled to a TOF MS 6224 mass spectrometer (Agilent Technologies), equipped with an ESI ion source. For the separation of metabolites, an Agilent Technologies fused silica capillary (total length, 96 cm; internal diameter, 50 µm) was used, working in normal polarity. Before each analysis, the capillary was washed for 5 min (950 mbar) with background electrolyte (BGE) (0.8 M formic acid solution in 10% methanol (v/v)). The sample injection was performed during 50 s at 50 mbar and, in order to improve the reproducibility of the analysis, the BGE was injected for 20 s at 100 mbar after the injection of each sample. The separation was performed with an internal pressure of 25 mbar at a voltage of +30 KV and at a constant temperature of 20 ° C. The total analysis time was 30 minutes. Mass spectrometry was operated in positive polarity, with a mass range 74–1000 m/z at a scanning speed of 1.00 spectrum /s. Other parameters fpr the MS were: fragmentor at 100 V, skimmer at 65 V, OCT RF Vpp at 750 V, drying gas temperature at 250 °C, flow at 10 L/min, nebulizer at 4 psig and capillary voltage at 4000 V. The sheath liquid used consisted of methanol:water (1:1, v/v), formic acid (1 mM) and two reference masses (5 µL of purine: 121.0509 and 15 µL of HP-0921: 922.0098) at a flow rate of 0.4 mL/min (1:100 of split ratio).

**Strengths and limitations of the study**

Our study's strengths include a unique cohort composed of participants from the European Prospective Study on Nutrition and Cancer (EPIC) followed for almost 15 years. The fact that none of the participants presented PD or disease-related symptoms at the sample collection time makes this study of exceptional value for discovering potential biomarkers for the early diagnosis of this devastating disease. The multiplatform approach adopted for this untargeted high-resolution metabolomics study shed light, for the first time, on the metabolic remodeling taking place many years before PD development in 33 plasma metabolites. Among those changes, five FFAs were detected as significantly affected by two independent analytical platforms (GC-MS and LC-MS), increasing the annotations' confidence and reinforcing the robustness of biological observations.

The study's limitations include a modest sample size, bestowing this work the category of a pilot study. Additionally, our cohort consists of subjects living in Spain, and, therefore, our findings may not be generalizable to other geographical areas. However, our results open up the door for future studies with more significant independent cohorts to replicate and validate such metabolites to consider them as prognostic biomarkers of PD development.

**Fig S1**. **PCA-X score plot for the 3 analytical platforms. A** ($R^2$ =0.396) represent the PCA-X model for LC-MS ESI (+) (UV scaling); **B** ($R^2$ =0.509) represent the PCA-X model for LC-MS ESI (-) (UV scaling); **C** ($R^2$ =0.356) represent the PCA-X model for CE-MS (UV scaling); and **D** ($R^2$ =0.691) represent the PCA-X model for GC-MS (Par-log scaling). The four models showed very good QC clustering, thereby indicating good system stability and reliability of the results (grey circles, pre-PD; blue squares, controls; green triangles, QCs).

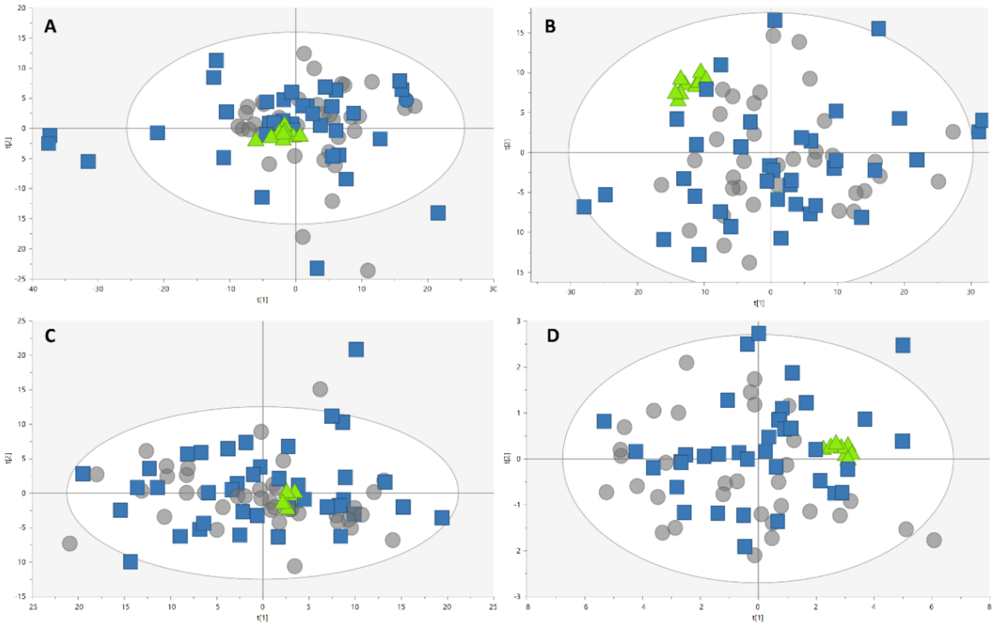


**Fig S2. Supervised OPLS-DA plot of LC-MS (+) data analysis**. The model presented good quality of variance explained and predicted variance ($R^2$ = 0.996, $Q^2$ = 0.687) (UV scaling), and a *p* CV-ANOVA of $1.31x10^{-8}$ (grey circles, pre-PD; blue squares, controls).

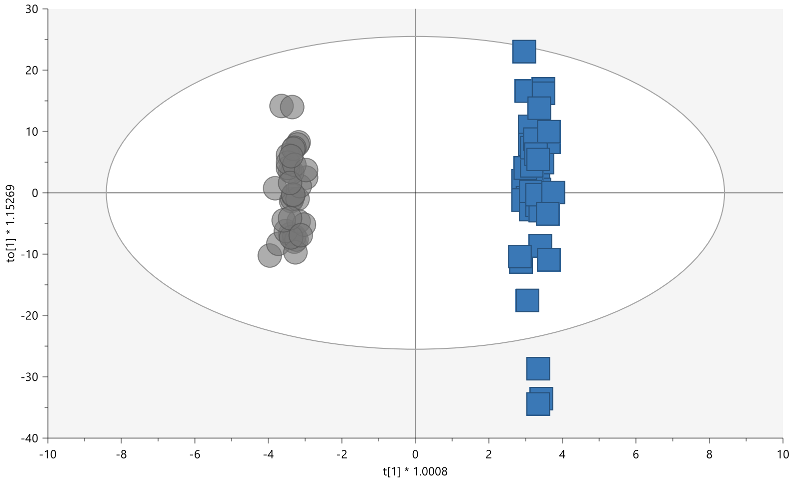

1. González, C.A.*, et al.* El estudio prospectivo europeo sobre cáncer y nutrición (EPIC)(#). *Revista española de salud pública* **78**, 167-176 (2004).
2. Riboli, E. & Kaaks, R. The EPIC project: rationale and study design. European Prospective Investigation into Cancer and Nutrition. *International journal of epidemiology* **26**, S6 (1997).
3. Riboli, E.*, et al.* European Prospective Investigation into Cancer and Nutrition (EPIC): study populations and data collection. *Public health nutrition* **5**, 1113-1124 (2002).
4. Godzien, J.*, et al.* Rapid and reliable identification of phospholipids for untargeted Metabolomics with LC–ESI–QTOF–MS/MS. *Journal of Proteome Research* **14**, 3204-3216 (2015).
5. Whiley, L., Godzien, J., Ruperez, F.J., Legido-Quigley, C. & Barbas, C. In-Vial Dual Extraction for Direct LC-MS Analysis of Plasma for Comprehensive and Highly Reproducible Metabolic Fingerprinting. *Analytical Chemistry* **84**, 5992-5999 (2012).
